# Implementing Inclusive Online Psychological Support after Stroke: Professional Stakeholder Views from a Qualitative Study

**DOI:** 10.1101/2025.06.21.25330050

**Authors:** Jasmine Ditta, Verity Longley, Kate Woodward-Nutt, Niki Chouliara, Shirley Thomas, Sarah Cotterill, Ann Bamford, Paul Conroy, Audrey Bowen, Emma Patchwood

## Abstract

**Background:** Emotional difficulties post-stroke are common, particularly among people from minoritised ethnic communities or with aphasia, an acquired communication disorder. However, there is a shortage of supportive evidence-based interventions, and of clinical psychologists to deliver them at scale. Wellbeing After Stroke-2 (WAterS-2) is a novel, online, group psychotherapy intervention based on Acceptance and Commitment Therapy. It was designed to support post-stroke psychological adjustment, delivered by a trained non-psychologist workforce to increase potential reach. To guide equitable implementation of such interventions into routine stroke care, this study explored barriers and facilitators from the perspectives of key stakeholders: healthcare commissioners, managers, and frontline clinicians.

**Methods:** Qualitative, semi-structured, individual interviews were conducted online with purposively recruited healthcare professionals involved in commissioning, managing, or delivering stroke care in England. The qualitative interview schedule was informed by both the Consolidated Framework for Implementation Research (CFIR) and Normalisation Process Theory (NPT), while the subsequent thematic template analysis was primarily guided by CFIR to explore contextual factors and practical strategies for equitable implementation.

**Results:** Fourteen interviews were conducted, with commissioning managers (*n* = 8), service managers (*n* = 4), and clinicians (*n* = 2). Analyses suggested three themes around the barriers and facilitators likely to influence equitable implementation: (1) Quality of Care vs. Quality of Cash Flow - highlighting a tension between tailoring for inclusion and demonstrating clinical- and cost-effectiveness; (2) Time & Leadership - highlighting a need for leadership to prioritise time investment in equity efforts; (3) Postcode Lottery - highlighting existing variation in local infrastructure, with third-sector partnerships suggested as key enablers.

**Conclusion:** Equitable implementation of online psychological support requires balancing the diverse needs of stroke survivors and the constraints of a publicly funded healthcare system. Recommended actions to enable equity provide useful insights for the development, implementation and commissioning of online mental health interventions for stroke survivors. Strategies such as embedding interventions within existing workflows, securing leadership support, and forming partnerships with community organisations may help translate online support like WAterS-2 into routine care.

## Background

Stroke is a leading cause of global disability (1). Approximately one-third of survivors experience post-stroke depression (2) and one quarter develop anxiety (3). These conditions are associated with increased mortality (4), slower rehabilitation (5) and reduced quality of life (6). In the UK, supporting psychological adjustment has been a top research priority for life after stroke since 2012 (7, 8). Yet, there remains a lack of evidence-based interventions and clinical psychologists to deliver them at scale (9). Consequently, stroke survivors often report feeling their psychological needs are unmet (10).

This gap is exacerbated by health inequalities, defined as “unfair and avoidable differences in health across the population and between different groups in society” (11). Health inequalities impact on the whole stroke pathway. For example, in the UK people from minoritised ethnic communities experience higher risk of stroke and poorer functional outcomes compared with White British people (12). Similarly, acquired communication problems such as aphasia are common post-stroke and result in poorer functional outcomes (13). These two examples of inequalities intersect with mental health post-stroke; people from minoritised ethnic communities and those with aphasia go on to experience disproportionately high mental health challenges post-stroke compared with the wider population (14, 15), and face barriers accessing appropriate mental health services (16). People from minoritised ethnic communities receive fewer referrals to mental health services compared with White British people (16), whilst many existing mental health interventions are not accessible for people with communication difficulties such as aphasia (17, 18). Therefore, there is a need for evidence-based psychological support in routine stroke care that is inclusive and accessible for people from minoritised ethnic communities and those with aphasia.

It is important to consider health equity beyond intervention development. Understanding how complex psychological interventions can be implemented can inform service improvement strategies (19). Previously, mental health interventions have faced challenges becoming integrated into routine care due to inadequate implementation planning (20, 21). Historically, stroke survivors from minoritised ethnic communities and those with aphasia have been underrepresented or excluded from research (22–24), meaning stroke care innovations are less well understood for these groups (18, 23, 25). If implementation processes do not consider health equity, these limitations may be inherited and perpetuated (26, 27). Early consideration of barriers and facilitators can inform design optimisations and prevent resource wastage on unsuccessful implementation (28, 29), enhancing the likelihood of equitable and successful implementation.

Acceptance and Commitment Therapy (ACT) combines mindfulness and acceptance strategies with behaviour therapy techniques. ACT enhances awareness, acceptance and understanding of one’s thoughts (30). It focuses on increasing psychological flexibility, helping individuals live meaningful lives, even amid adversity (30, 31). There is growing evidence supporting ACT’s efficacy in improving psychological wellbeing and preventing depression post-stroke (32–35). Participants report finding ACT beneficial in fostering acceptance of an altered reality and adapting to stroke-related disabilities (35). Furthermore, the latest National Clinical Guideline for Stroke for the UK and Ireland (36) recommended ACT for stroke survivors with anxiety or depression. Despite growing evidence, delivering this at scale to the diverse stroke population remains a challenge.

Previously our team co-developed the Wellbeing After Stroke (WAterS) intervention with stroke survivors, carers and healthcare professionals (37). WAterS is a novel, online ACT-informed psychotherapy group intervention; designed to reduce distress and improve post-stroke psychological adjustment. Co-Development included Patient Carer and Public Involvement (PCPI) and a commitment to inclusivity and accessibility, with consideration of aphasia. Additionally, the intervention includes an online staff training programme, delivered by a clinical psychologist, to equip staff without prior psychological expertise with the skills to deliver the intervention. After development, WAterS was tested in a feasibility study (37). This study indicated the feasibility of remotely training a third-sector / (charity) workforce to deliver WAterS online to groups of stroke survivors. The study was carried out within one UK charity, which may not fully capture the diversity of practices and challenges encountered in different healthcare environments. Subsequently we enhanced the intervention through more PCPI co-development. This included further consideration of how the intervention might be adapted for implementation in more settings; such as within the UK National Health Service (NHS), and delivered to under-served communities, beginning with two: minoritised ethnic groups and people with aphasia. We refer to this intervention as WAterS-2 (38).

Online delivery in stroke care has the potential to reduce inequalities in access by improving service reach in a resource efficient way (23, 39). Additionally, psychological telehealth interventions have been found feasible to deliver, with stroke survivors finding them acceptable (40), accessible and helpful (41, 42). There is limited research on how to equitably implement online mental health interventions like WAterS-2 for stroke survivors into routine care. A 2024 systematic review (43) identified barriers and facilitators to implementing online interventions in mental health care in any setting. This review suggested that both patients and providers perceive digital technologies as an additional burden (43). Yet, recent research indicates that online delivery may be more convenient for stroke survivors, due to common mobility impairments that make in-person appointments more difficult (44). Additionally, health inequalities may become exacerbated if not accounted for during implementation. Online mental health interventions need to be adapted to meet the needs of users from different backgrounds, including different ethnic groups otherwise lack of cultural sensitivity poses a barrier to implementation (43).

This paper aimed to understand the implementation context by identifying the beliefs held by healthcare commissioners, managers, and clinicians regarding the barriers and facilitators to the equitable implementation of an intervention like WAterS-2 into NHS stroke care.

## Method

### Design

This study employed individual semi-structured qualitative interviews. A subtle realist approach (45) was adopted, which acknowledges the researcher’s perspective is shaped by their social world, but also that phenomena can exist independently. This was chosen to recognise the subjectivity of participants and researchers while also assuming findings have the potential to have wider relevance. Ethics approval was granted by the University of Manchester Research Ethics Committee (UREC; Reference: 2023-18486-32352; Date: 21/12/2023). Reporting adhered to the consolidated criteria for reporting qualitative research (COREQ) (46).

### Participants

Professional stakeholders involved in stroke care were purposively recruited from three groups: commissioning managers, service managers, and clinicians. An email was sent to operational delivery networks and research networks, including individuals and organisations from research institutions, clinical settings, and third-sector organisations (e.g., charities). Contacts were asked to distribute an advertisement poster to their relevant networks, including via social media posts on Twitter / X page e.g. via (@StrokeWaters). A snowball sampling method was also employed to identify participants.

The eligibility criteria were:

1. Adults aged 18 years or older.
2. Current/previous experience delivering, commissioning or managing UK stroke care.
3. Knowledge of the NHS healthcare system and implementation of interventions.
4. Capable of providing informed consent.
5. Willing and able to engage in an online interview conducted in English.

Upon expressing interest, potential participants were assessed for eligibility by EP through email or telephone discussions. Eligible individuals received the Participant Information Sheet, detailing the study’s purpose, procedures, and potential risks. Interviews were arranged at a time of the participant’s convenience and an orienting document was sent in advance to promote understanding and maximise explorations (see Appendix A for orienting document)

### Materials

The interview schedule was informed by the Consolidation Framework for Implementation Research (CFIR) and Normalisation Process Theory (NPT) (47, 48) CFIR’s meta-theoretical framework guided the systematic appraisal of implementation contexts, including potential barriers and facilitators (47). NPT aided understanding of how people think about, use and sustain new innovations (48).

The interview schedule incorporated 13 open-ended questions with probes related to the equitable implementation of WAterS-2. Interview questions were mapped onto both CFIR and NPT domains to understand context and practical strategies that would inform implementation (See Appendix B for ‘mapped’ interview schedule)

Participants were asked to focus particularly on issues for stroke survivors from minoritised ethnic communities and people with aphasia, due to their increased risk of mental health problems post-stroke. For example: *What evidence or guidelines do you look for to commission / adopt services or interventions, such as WAterS-2? Tell me about any strategies you already use to support minoritised ethnic communities in accessing and using post-stroke services? How do influential voices and leadership within your organisation influence the adoption of new interventions?*

### Data Collection

All one-off interviews were conducted by a female member of the research team; EP (research fellow), with over 15 years of experience of qualitative interviewing. Interviews were online (via Zoom or Microsoft Teams) and recorded. After each interview EP recorded field notes, reflecting on the context, researcher-respondent interaction, and initial thoughts about the interview (49). The number of interviews was determined by content saturation, with an anticipated maximum of 15 (50). Interviews were transcribed using Zoom / Microsoft Teams, then checked by reviewing recordings alongside the transcripts to incorporate non-verbal cues and ensure meaning was appropriately? captured. Identifiable information (e.g., names, places, and dates) was replaced with generic terms. Each participant was assigned a number. A separate document linking these pseudonyms to original identities was created and stored separately. Transcripts were not returned to participants for comment or correction.

### Data Analysis

Data were managed using NVivo version 12 (51). Data analysis was conducted by JD, with input from EP and KWN. Findings were thematically analysed using template analysis (52, 53), a systematic method which created a hierarchical coding “template”, allowing exploration of the research questions using CFIR constructs as *a priori* themes while incorporating inductive analysis to allow for unanticipated themes from the data (54). Analysis followed these steps (53):

1. Familiarisation with data, achieved through cleaning transcripts and reading without notetaking.
2. Initial themes derived from a subset of the data, focusing on recurring ideas, aspects relevant to the research question and the detailed CFIR constructs as *a priori* themes.
3. Themes organised into meaningful hierarchical groups, defining how they relate to each other, the dataset and the research objective. A-priori themes were revised or discarded if they proved ineffective to capture key meanings in the data.
4. An initial template was constructed from the themes identified from a subset of the first seven interviews, chosen for a variety of participant roles and viewpoints.
5. The initial template was applied to full dataset by coding each transcript, adjusting the template as appropriate by refining, adding or removing themes.
6. Themes were presented to EP and discussed to facilitate reflections on analysis.
7. The template was finalised through, incorporating feedback from EP, re-evaluating, merging, splitting, and redefining themes to improve clarity and relevance.
8. The finalised template was applied across the entire dataset.

While presented sequentially, analysis was an iterative process, involving continual revisiting and revising of steps.

### Validity and Reflexivity

Approaches to enhance validity were aligned with a subtle realist position (45). Critical evaluation and reflection on any unrecognised assumptions were encouraged through reviews on analysis by EP (54). However, the reliability of these reviews was not assessed, as this position acknowledges and values researcher subjectivity in knowledge construction (55, 56). Additionally, a reflexive diary documented JD’s influence on interpretation, considering their roles as a support worker for individuals with brain injury, a novice researcher and a person from an ethnic minority group. New insights were gained by revisiting this reflexive space from JD and EP’s perspectives, enriching analysis.

## Results

### Participants

Fourteen interviews were conducted between February 2024 and April 2024, including commissioning managers (*n* = 8), service managers (*n* = 5) from NHS and charitable organisations, and clinicians (*n* = 2, occupational therapist and nurse). There were 10 females and four males, with a median age of 51 years old (range 26 to 56) and a median experience of 11.5 years (range: 1.5 - 30 years). Twelve participants identified as White British, two as Black/Black British/Caribbean/African, and one as Mixed/Multiple ethnicity. Interviews lasted between 33 and 79 minutes (mean 56).

### Findings

Analysis resulted in three themes influencing the barriers and enables to the equitable implementation of online group psychological support interventions: ‘Quality of Care v Quality of Cash flow’, ‘Time & Leadership’, and ‘Postcode Lottery’.

#### Theme 1: Quality of Care v Quality of Cash flow

All participants believed that current emotional support for stroke survivors is inadequate, highlighting the need for interventions like WAterS-2 to better meet survivors’ needs: “*We’re desperately short of strategies to help people in this situation” (P01, service manager),* Participants emphasised that psychological support should be available to all stroke survivors, highlighting a moral imperative to ensure inclusive care. When asked how this can be achieved for survivors from minoritised ethnic groups or with aphasia it was suggested that interventions be tailored to the needs of target groups:

> “what’s worked best, is where there’s been a really targeted effort on outreach and on ensuring that services are appropriately responsive to any specific needs of a minoritised group” (*P11, commissioning manager*).

Participants discussed ideas for how this approach could be applied to interventions like WAterS-2, reflecting on past experiences. For example, targeting sessions exclusively for specific minoritised ethnic groups, delivered by staff from the same background. Participants from minoritised ethnic backgrounds themselves spoke of stigma and stereotypes that can impact on help-seeking, suggesting they could better understand the barriers for patients from minoritised ethnic groups. Some participants emphasised the importance of including these voices in intervention delivery and design:

> “the advantage to being a black person…I might have gone through some of what this person is talking about. I might know some of these barriers already. But because I am now trained personnel, I am able to use my knowledge of what that person might be going through with what I have” (*P14, commissioning manager*)

For those with post-stroke aphasia, participants suggested adapting the content: “you could say… the session is shorter, it is slower, it’s only focused on one theme” (*P04, service manager*). Some participants had experience of tailoring services to people with aphasia and found these to be “*well received”* (*P14, commissioning manager)*. However, there appears to be a tension balancing this customisation with commissioning realities. There was an assumption that targeted interventions use more time and resources and this may impact cost-effectiveness. Participants in commissioning roles indicated that for WAterS-2 to receive funding, it needs to demonstrate effectiveness in improving patient outcomes and cost-efficiency.

> “From a sort of commissioning point of view…we’re interested in deploying things that work, and have an impact, and are cost effective” (*P11, commissioning manager*)

Service managers and clinicians suggested that in their experience of applying for funding, costs and cost-effectiveness often overshadow patient need:

> “the whole conversation isn’t really about quality of care, it’s *actually* all about money” (*P07, service manager*).

Therefore, service managers and clinicians suggested that broadening the scope of psychological support would increase the likelihood of securing funding, although the impact this would have on inclusivity (e.g. people from minoritised ethnic communities or with aphasia) was not always discussed or unpicked within the interview context. For example, combining stroke and neurological populations in community NHS teams or integrated interventions that addresses numerous conditions, demonstrates how broad applicability can streamline implementation across a larger patient population. Such approaches appealed to commissioners:

> “If this could be applicable to acceptance for people in neuro… that would be sustainable. You’re reaching more people” (*P08, commissioning manager*)

In summary, tailoring interventions was seen as key to increasing inclusivity for minoritised ethnic communities and people with aphasia. However, participants highlighted a trade-off between tailoring and demonstrating cost-effectiveness, with broader, cross-condition approaches perceived as more scalable. This reveals a core tension between designing for equity and aligning with commissioning priorities in a resource-constrained system.

#### Theme 2: Time & Leadership

Participants indicated that achieving equitable implementation requires an investment of time. For example, it was suggested that healthcare professionals should practice cultural competence by taking the time to understand patients’ cultural backgrounds and how this shapes their care needs:

> ‘It’s not a sort of, you know, oh I understand what it means to be from this particular minority group, it’s about understanding your role in making cultural adaptations, and being curious about that, and being prepared to talk about it’ (*P11, commissioning manager*).

Equally, whilst telehealth interventions can be time-saving, they require investment of time to support patients to use them. Participants reported assumptions that limited technological skills and socioeconomic circumstances would pose a potential barrier to implementation:

> ‘‘typically in [location]…you’ve got quite an older population that might struggle to access online services and then we’ve also got quite a younger population who are quite socially deprived who can’t even use a smartphone” (*P07, service manager*)

Participants emphasised that dedicating time to assist individuals in using online services such as by providing instructions, increased inclusivity:

“It took more of our time to ensure that we included people. But we needed to do that because you cannot exclude them” (*P14, commissioning manager*)

However, balancing these efforts with the demands of a busy healthcare setting appears to be challenging. Participants reported that heavy workloads often left little time to learn and implement new interventions:

> “at the moment people are firefighting, papering over the cracks” (*P07, service manager*)

Even instances when time and funding were allocated, these resources were sometimes not used due to this heavy workload:

> “you could have kind of like 5 days in your year where you could go for funded training… but we usually just go and work on our caseload instead” (*P06*, *clinician*)

This suggests that the additional time needed for inclusivity efforts may hinder implementation unless it could be prioritised. Despite this, it appears that time-efficient online interventions may be popular in the context of routine stroke care. For example, participants supported the potential value of WAterS-2 for its ability “*to hit more people” (P07, service manager)*, being a more *“efficient use of time” (P01, service manager)* and providing “*standardisation” (P05, service manager).* Participants suggested that having a senior member of staff to lead or encourage implementation can motivate the rest of the team to dedicate time towards it:

> “if a clinical psychologist in your team says, this is great intervention we need to be providing it - how are we going to go about that? *That’s* going to get your buy-in of the community team… which is what happened with [Training Program], we got 80 people signed up” (P07, service manager)

In summary, whilst time is essential for promoting equity - through cultural competence and digital inclusion - workload pressures often limit its availability. Broadly-speaking, having clinical champions as leaders to advocate for intervention implementation was suggested to foster broader team buy-in and encourage allocation of time toward training.

#### Theme 3: Postcode Lottery

Many participants suggested that embedding psychological interventions like WAterS-2 into existing workflows could facilitate implementation. This approach was seen to reduce the cost and workload that might be associated with a separate, standalone initiative:

> “it’s more efficient to have it delivered as part of a psychology that’s already there” (*P08, commissioning manager)*

However, there appears to be significant resource variability across settings, which means some locations lacked the necessary infrastructure into which new interventions could be embedded, if pre-existing psychological support was a requirement. For example, some participants highlighted that their stroke services currently offer limited or no psychological support:

> “We’ve had no psychologists specifically for stroke for about 10 / 11 years” *(P05, service manager)*

Despite WAterS-2 being designed for delivery by a non-psychologist workforce, this lack of existing infrastructure may make implementing interventions like WAterS-2 more costly and logistically challenging when considering training and supervision requirements. Therefore, while embedding psychological interventions into existing workflows can facilitate implementation for some organisations, this approach may not be feasible for more resource-limited settings. However, collaborating with third-sector organisations was suggested as potentially offering a solution. Many participants highlighted the value of *“joint work between the NHS and voluntary sector” (P12, commissioning manager)* to extend psychological support to stroke survivors, especially in settings lacking these services.

Therefore, participants suggested that third-sector partnerships could expand access to interventions like WAterS-2 in under-resourced areas. It was also suggested that collaborating with third-sector organisations could enhance access for minoritised ethnic communities. These organisations often have established connections within communities that can be leveraged to reach underserved groups:

> ‘you expect people to kind of turn up in *your* environment as an NHS provider. And actually, not everybody feels particularly comfortable with that… so doing sessions with groups that already exist; that are centred around those populations with their cultural differences’(*P07, service manager*)

Therefore, in summary, a tension emerged: while psychological support was viewed as essential everywhere, participants also felt implementation would be easier where infrastructure already exists. This highlights a paradox—those most in need may face the greatest barriers unless deliberate action is taken to close the gap. Partnering with third-sector organisations was seen as a key way to improve access, especially in underserved areas.

## Discussion

This qualitative study explored barriers and facilitators to equitably implementing an intervention like WAterS-2; drawing on insights from healthcare commissioners, managers, and clinicians. While participants supported the intervention’s aims—particularly its potential to address gaps in support for underserved stroke survivors—they also highlighted significant systemic constraints. Three themes were developed: (1) the tension between tailoring for equity and the need for scalable, cost-effective models; (2) the challenge of time and capacity, highlighting a need for leadership to prioritise time investment in equity efforts since limited resources impede implementation despite goodwill; and (3) the influence of local context, revealing a ‘postcode lottery’ in psychological stroke care and highlighting a paradox—those most in need may face the greatest barriers unless deliberate action is taken to close the gap. It was suggested that third-sector partnerships and flexible delivery models could be potential solutions. Overall, findings emphasise the need to balance accessibility, sustainability, and clinical quality in efforts to embed inclusive interventions like WAterS-2 into routine care.

Participants highlighted the need for inclusive interventions to better address the psychological needs of stroke survivors. Tailored approaches were suggested, such as targeted sessions specifically for minoritised ethnic groups, and/or led by staff from the same background. Evidence suggests that treatment provided by same-ethnicity patients and therapists in mental health services is preferred by patients, improves adherence, therapeutic relationships, and patient satisfaction however has minimal impact on treatment outcomes (57, 58). Similarly, whilst our interviews tended to focus more on inclusion of minoritised ethnic groups, participants also suggested having aphasia-specific groups, with shorter, slower-paced sessions for this participant demographic. Previous research has suggested that post-stroke aphasia support groups foster a sense of belonging through mutual understanding and shared experiences (59, 60). However, specialist knowledge and training is needed to effectively deliver aphasia-specific psychological support (61). Furthermore, individuals with post-stroke aphasia can struggle to engage with general group-based programmes (62). Offering smaller, aphasia-specific groups inclusion of non-verbal activities could improve engagement and therapeutic outcomes among these individuals.

Our findings suggest a tension between tailoring interventions and the cost of inclusion. While commissioners suggested they value patient need, service managers and clinicians implied that cost-effectiveness often takes precedence during funding decisions. This illuminates the pragmatic realities of publicly funded healthcare systems, where economic imperatives can overshadow clinical ideals and patient need. While tailoring interventions to address health inequalities may offer promising benefits in patient care, it requires balancing quality of care and cost-effectiveness. In this light, participants proposed broadening interventions like WAterS-2 to include neurological populations to increase funding prospects. Although research on what informs healthcare commissioners’ decisions is limited, it is increasingly common for the NHS to combine stroke and neurological services (63–65), suggesting a trend toward broader service integration. ACT has been used in mixed neurological populations (66), however WAterS-2 was in fact developed to tailor ACT to stroke specifically. Therefore, while expanding the scope of an intervention like WAterS-2 may facilitate implementation, this approach risks exacerbating existing health inequalities by potentially overlooking the unique needs of stroke survivors.

Participants suggested a need for healthcare professionals to invest time to ensure inclusivity, for example through cultural competence training to promote equitable care, or technical training to overcome barriers related to access to technology or digital skills (67, 68). While cultural competency and digital skills training can promote equitable access, our findings suggest that the additional time required for these efforts may hinder implementation. Time constraints have consistently been identified as a barrier to implementing *any* healthcare intervention (69–71). However, participants suggest that clinical champions or leaders could drive implementation. Clinical champions are a widely used strategy to reduce barriers (72, 73) and facilitate implementation by providing knowledge, fostering teamwork and cultivating a learning culture (74). These skills could potentially be leveraged to enhance the delivery of equitable services.

Lastly, it was suggested that embedding interventions within existing workflows can facilitate implementation. Previous literature indicates interventions integrated into existing practices demonstrate greater adherence and sustainability (75). However, there appears to be significant variability in resources across stroke services, with some lacking the necessary infrastructure for integration. Some participants reported collaborating with third-sector organisations to provide psychological support to stroke survivors and to reach minoritised ethnic communities. Collaborations between sectors to improve healthcare access and optimise resources are not novel (e.g., hospitals partnering with faith-based organisations to increase vaccination rates (76)). However, the use of such collaborations for delivering mental health interventions is less established. The WAterS2 flexible training model, which does not require prior psychological expertise and utilised online platforms to deliver, enables its implementation beyond clinical settings and geographical catchment areas. Research suggests that these community organisations are often particularly client-led, adept at reaching marginalised populations, and perceived as more approachable than traditional healthcare providers (77, 78). Additionally, while these organisations are eager to be more evidence-informed, they often face limitations in staff skills and resources (79, 80). Therefore, such partnerships could make interventions like WAterS-2 more accessible and potentially reduce health inequalities.

The study had strengths and limitations. First, online interviews enabled a broad geographic reach, capturing diverse perspectives across England. While online interviews may lack some non-verbal cues associated with in-person interactions (81), video recordings captured these cues and were incorporated into the transcripts. This helped maintain the depth of understanding. Including commissioning managers, service managers, and frontline clinicians brought unique insights reflective of each group’s distinct roles and experiences. This diversity of viewpoints provided a more nuanced understanding of implementation. However, the predominantly White British sample presented a limitation. While largely representative of the UK healthcare workforce (82), given the focus of the study it may have limited the depth of insights. For example, participants from minoritised ethnic communities were able to provide beneficial insights into enhancing inclusivity and an insider perspective into why certain strategies may be effective, however we did not explore this in depth.

The use of CFIR and NPT (47, 48) theories presented opportunities and challenges. Use of theoretical frameworks during data collection facilitated a comprehensive investigation of potential implementation barriers and facilitators. However, during analysis, the general focus of theory may not have suited capturing the unique contextual challenges identified. While CFIR has been recommended for examining equity and implementation (83), studies have noted limitations in addressing equity-related complexities (84, 85). Future research could potentially benefit from integrating equity or justice theories alongside implementation theories like CFIR and NPT, enabling a more comprehensive understanding of implementation dynamics.

A strength was the use of template analysis. Unlike other thematic approaches (e.g., Framework Analysis), template analysis does not impose a predefined sequence of coding levels (54). Instead, it encourages the development of themes more extensively where the richest and most relevant data to the research question are found. This flexibility proved valuable in capturing the nuanced complexities during analysis. Instead of rigidly adhering to a structured coding framework, for example, template analysis allowed for a more iterative and exploratory approach, yielding richer insights into the research question, which suited the study’s exploratory nature. Another strength was employing reflexivity. Independent reviews of analysis enhanced awareness of researcher subjectivity and promoted critical analysis. Although time-consuming, reflexivity enhances analysis quality and contributes to more robust knowledge in the field (86).

Overall, findings provide valuable insights for stakeholders involved in developing, implementing and commissioning online mental health services in stroke care, particularly those that are group-based and/or ACT-informed. They underscore the complexity of implementing psychological support into routine care. Based on findings, the following recommendations are proposed to facilitate equitable implementation:

1. **Demonstrate Cost-Effectiveness**: Highlight the economic value of interventions, particularly the cost-benefits of reaching marginalised groups, such as those from minoritised ethnic communities and people with aphasia. This approach can gain stakeholders’ support and facilitate commissioning and integration into everyday practice.
2. **Prioritise Leadership and Advocacy**: Target strategic clinical and managerial leaders to enhance their knowledge, skills, and motivation to support equitable implementation.
3. **Partner with third-sector organisations:** Assess organisational resources and opportunities for collaboration with third-sector organisations to facilitate implementation.

## Supporting information

Appendix A - Orienting information for interviewees

Appendix B - 'mapped' interview schedule

## Data Availability

All data produced in the present study may be made available upon reasonable request to the authors

## Abbreviations

ACT: Acceptance and commitment therapy
WAterS: Wellbeing After Stroke
CFIR: Consolidation Framework for Implementation Research
NPT: Normalisation Process Theory

## Declarations

### Ethics approval and consent to participate

This project was assessed and approved by the University of Manchester Research Ethics Committee (UREC; Reference: 2023-18486-32352; Date: 21/12/2023). Informed consent was obtained from all individual participants included in the study. All interviews were performed in accordance with relevant guidelines and regulations.

### Consent for Publication

Not applicable.

### Availability of data and materials

The datasets used and analysed during the current study may be made available from the corresponding author on reasonable request.

### Authors’ contributions

All co-authors contributed to the study’s conception and design, material preparation and writeup. EP performed data collection. JD preformed analysis with input from EP and KWN. The first draft of the manuscript was written by JD as part of an MSc dissertation project, with AB as primary supervisor and EP as secondary supervisory.

## Acknowledgements

We would like to thank research participants who contributed their time for interviews. We would also like to thank all members of the WAterS and WAterS-2 research team, including the Study Management Group, PCPI Research Advisory Panel, and Study Steering Committee. Finally, thanks to Stroke Association who funded the project (project grant PG22/23_S2100010)

