## Appendix A - Orienting information for interviewees for "Implementing Inclusive Online Psychological Support after Stroke: Professional Stakeholder Views from a Qualitative Study"

**Appendix A Orienting information sent to interviewees in advance**

**Wellbeing after Stroke-2 (WAterS-2)**

**Interviews to explore *how* we could implement this intervention in practice**

### 1. What is WAterS-2?

**It aims to promote psychological adjustment after stroke and prevent mental health crises**

- Stroke Survivors are eligible for intervention if they are 4 months or more post-stroke and have had difficulties adjusting, or are at risk of developing mental health issues.
- The intervention is for **groups** (4 to 8 stroke survivors per group) and:
  - Delivered **Online** (e.g. videoconferencing) with 9 sessions over 9 consecutive weeks.
  - Informed by **Acceptance and Commitment Therapy (ACT)** using **structured session plans** with 'scripts' to help staff deliver consistently.
  - Each of the 9 weekly sessions requires approximately **2.5 hours** allotted time for staff
  - Each session is facilitated by **two staff** (one who 'leads' and one who 'supports')
- Training programme for facilitators:
  - Initial Training: **4 x 3.5 hour online sessions** over 4 consecutive weeks, delivered **online** by a Clinical Psychologist
  - Support available: 1 hour per week. Online group supervision with a Clinical Psychologist
  - Facilitators do not need previous experience with Acceptance and Commitment Therapy. They would need:
    - Experience / knowledge of stroke and ideally of delivering wellbeing support;
    - Experience of facilitating groups; ideally using online tech / videoconferencing
    - The willingness / motivation and time to participate (see above). Minimum time required is for initial training and then for facilitating one group.

### 2. What is the WAterS-2 research project?

**WAterS-2 has been funded by the Stroke Association (2023-2025) to explore:**

- How can we identify, train and support a **wider workforce** to facilitate WAterS, including NHS-funded staff (previously, WAterS has been delivered by Stroke Association staff).
- How can we ensure WAterS-2 is available to everyone; whilst *particularly* thinking about:
  - Stroke survivors from **minoritized ethnic communities**?
  - People with **communication disabilities**?

The interview will explore your opinions on implementing WAterS-2 in practice. For example: Could it fit into existing commissioning and workloads? What staff could be trained to facilitate? How might we reach stroke survivors? How should we measure 'success'?

And more….

We want your honest opinions: don't hold back! Everything will be confidential.

We look forward to speaking with you.
