## Appendix B - 'mapped' interview schedule for "Implementing Inclusive Online Psychological Support after Stroke: Professional Stakeholder Views from a Qualitative Study"

**Appendix B**. Interview Schedule – mapped to CFIR and NPT Domains

Questions were informed by both the Consolidated Framework for Implementation Research (CFIR) (Nevedal et al, 2021) and Normalization Process Theory (NPT) (May, 2009; Murray, 2010).

The table below summarises the questions (and probes) for each participant type and how they map onto CFIR / NPT domains:

- Group 1: Commissioning managers
- Group 2: Service managers
- Group 3: Clinicians

| | | **Asked of Group** | | | | |
|---|---|---|---|---|---|---|
| **Question** | **Probes** | **1** | **2** | **3** | **CFIR Domain(s)** | **NPT Construct(s)** |
| WAterS-2 is about offering inclusive, accessible support to help people psychologically adjust to stroke and *prevent* a mental health crisis. How does this align with your commissioning goals? | Could you tell me about any services that are commissioned across geographical boundaries?<br><br>If not a good fit, explore alternative services that may be better suited? | ✓ | ✓ | | • Intervention Characteristics<br>• Outer **and** Inner Setting;<br>• Characteristics of Individuals | • Coherence<br>• Collective Action |
| What factors influence the adoption of new services in your area? | What evidence or guidelines do you look for to commission / adopt services or interventions, such as WAterS-2?<br><br>How do influential voices and leadership within your organisation influence the adoption of new interventions?<br><br>Resource / cost considerations and how they are calculated? | ✓ | ✓ | | • Process<br>• Outer **and** Inner Setting<br>• Intervention characteristics | • Collective Action<br>• Coherence |

| **Question** | **Probes** | **1** | **2** | **3** | **CFIR Domain(s)** | **NPT Construct(s)** |
|---|---|---|---|---|---|---|
| Can you tell me about any instances when you have commissioned/ provided interventions targeted at minoritized ethnic groups? | Do you have any thoughts on how interventions are made suitable for these groups and/or any suggestions for us when seeking expertise in including minoritized ethnic communities locally?<br><br>Are there existing initiatives focused on equity and inclusion that could apply to WAterS-2? | ✓ | ✓ | | • Process<br>• Outer **and** Inner Setting | • Collective Action<br>• Cognitive Participation |
| Tell me about any strategies you already use to support minoritized ethnic communities in accessing and using post-stroke services? | Do you have any thoughts on how interventions are made suitable for these groups and/or any suggestions for us when seeking expertise in including minoritized ethnic communities locally?<br><br>Are there existing initiatives focused on equity and inclusion that could apply to WAterS-2? | | ✓ | ✓ | • Outer setting<br>• Process<br>• Characteristics of Individuals | • Cognitive Participation<br>• Collective Action |
| What about strategies for supporting people with aphasia to access? | (same prompts) | | ✓ | ✓ | • Process<br>• Characteristics of Individuals | • Cognitive Participation<br>• Collective Action |
| If you implemented WAterS, what information would you want to collect to evaluate its success? | Any key performance indicators we should be aware of?<br><br>What would 'success' look like, in your opinion? | ✓ | ✓ | | • Intervention characteristics<br>• Process | • Reflexive Monitoring |

| Question | Probes | 1 | 2 | 3 | • CFIR Domain(s) | • NPT Construct(s) |
|---|---|---|---|---|---|---|
| Considering the information received prior to the interview, which frontline staff might be best positioned to deliver WAterS-2? | Do they know about Acceptance and Commitment Therapy?<br><br>How is staff time currently allocated? How do you release and compensate staff for training and intervention delivery?<br><br>What might motivate staff to be part of WAterS-2? | | ✓ | ✓ | • Inner Setting | • Collective Action |
| If you implemented WAterS-2, how would you identify suitable stroke survivors to receive? | Are there differences for reaching people from different demographics (e.g., minoritized ethnic communities / people with aphasia)? | | ✓ | ✓ | • Process | • Cognitive Participation<br>• Collective Action |
| What experience does your organisation have with videoconferencing tools like Zoom or Teams for staff training and service provision? | How do you find using remote technologies? | | ✓ | ✓ | • Inner setting | • Collective Action |
| Overall, what do you think are the main challenges to delivering WAterS-2? | How might these barriers be mitigated? | ✓ | ✓ | ✓ | *Answer dependent? Could be all* | *Answer dependent? Could be all* |
| Overall, what do you think could be the biggest enablers for delivering WAterS-2 in your service? | | ✓ | ✓ | ✓ | *Answer dependent? Could be all* | *Answer dependent? Could be all* |

| Question | Probes | 1 | 2 | 3 | • CFIR Domain(s) | • NPT Construct(s) |
|---|---|---|---|---|---|---|
| WAterS-2 is informed by Acceptance and Commitment Therapy. Do you or your colleagues have much experience with this therapy? | What further information would you want to receive to help make an informed decision about delivering WAterS-2?<br><br>What further training or support might you want to deliver WAterS-2? | | | ✓ | • Characteristics of Individuals | • Collective Action |
| Is there anything else you'd like to say about implementing WAterS-2? | | ✓ | ✓ | ✓ | n/a | n/a |
